# Spatially fractionated stereotactic body radiotherapy (Lattice SBRT) for large tumors

**DOI:** 10.1101/2020.03.09.20033332

**Authors:** Sai Duriseti, James Kavanaugh, Sreekrishna Goddu, Alex Price, Nels Knutson, Francisco Reynoso, Jeff Michalski, Sasa Mutic, Clifford Robinson, Matthew B Spraker

## Abstract

Stereotactic body radiotherapy (SBRT) has demonstrated clinical benefit for patients with metastatic and/or unresectable cancer. Technical considerations of treatment delivery and sensitive organs at risk (OARs) limit the use of SBRT in large tumors or those in unfavorable locations. Spatially fractionated radiotherapy (SFRT) delivers high-dose radiation to discrete sub-volume vertices within a tumor target while restricting the remainder of the target to low dose. SFRT has been utilized for treatment of large tumors with reported dramatic tumor response and minimal side effects. Lattice is a modern approach to SFRT that can be delivered with arc-based therapy, which allows for the rapid dose fall-off required for high quality SBRT. In order to overcome the limitations of SBRT for large tumors, we developed Lattice SBRT. Here we report the results of a dosimetry and quality assurance (QA) feasibility study of Lattice SBRT in 11 patients with 12 tumor targets, each ≥ 10 cm in an axial dimension. Prior CT simulation scans were used to generate volumetric-modulated arc therapy (VMAT) Lattice SBRT plans that were then delivered on clinically available Linacs. QA testing included external portal imaging device (EPID) and ion chamber (IC) analysis. All generated plans were able to meet the standard SBRT dose constraints, such as those from AAPM Task Group 101. Additionally, we provide a step-by-step approach for generating and delivering Lattice SBRT plans using commercially available treatment technology. Lattice SBRT is currently being tested in a prospective trial for patients with metastatic cancer needing palliation of large tumors (NCTXXXX).

## Introduction

Metastatic or unresectable cancer is responsible for significant morbidity and mortality in patients with solid tumors ^1^. SBRT is emerging as a high-value treatment option for such patients, offering improved symptom palliation across cancer types and even extended survival in oligometastatic populations. Unfortunately, SBRT can be difficult to deliver safely for large tumors. Prior studies have shown that SBRT may be associated with unacceptable toxicity for tumors greater than 5 cm ^2,3^. Additionally, large tumors may be near surrounding OARs, which can make planning difficult ^4,5^.

SFRT is a radiotherapy technique that theoretically allows for safe dose escalation for large tumors. Specialized beam collimation creates high-dose “peaks” organized throughout a target volume with intervening low-dose “valleys” ^6^. SFRT planned with two-dimensional techniques, such as GRID, has been evaluated for large soft tissue sarcomas, and is associated with excellent local control and low toxicity in prior case series ^7,8^. The GRID technique attempts to achieve a differential high-dose “peak” surrounded by lower-dose “valleys” by use of either a pre-cast block or multi-leaf collimators (MLCs) ^9^. While GRID is more widely-accessible, newer SFRT techniques, such as Lattice, which creates high-dose “islands” within a “sea” of lower dose, offer improved dose distribution and OAR sparing as compared to GRID, which may be beneficial for large or deep-seated tumors^10^.

In order to safely deliver SBRT to large tumors, we created a Lattice SBRT technique that delivers 2000 cGy in 5 fractions to the entire tumor target with a simultaneous integrated boost (SIB) of 6670 cGy to vertices arranged geometrically inside the tumor. Here we describe the Lattice SBRT planning process, resulting dosimetric parameters of 12 pilot Lattice SBRT plans, and their QA results.

## Materials & Methods

This study was approved by our institutional IRB. We retrospectively identified 12 large tumors greater than 10 cm in 11 patients previously treated at our institution. Patient and tumor characteristics were extracted from the electronic medical record (EMR).

A step-by-step guide to the Lattice SBRT contouring and treatment planning process is available in the supplementary materials (Supplement 1). The Lattice SBRT prescription was created on the assumption that the tumor planning target volume (PTV) should receive at least a standard 5-fraction palliative dose of 2000 cGy. Spatially fractionated techniques have traditionally created a peak-valley dose gradient of approximately 100%-30% ^6^, therefore a SIB of 6670 cGy was selected as a Lattice boost dose prescription.

To generate the desired high gradient dose distribution, our Lattice SBRT technique utilizes a geometric arrangement of spherical vertices, each with a diameter of 1.5 cm, a 6 cm center-to-center spacing, and a separation of 3.0 cm between each successive axial plane of spheres. A representative schematic is shown in Figure 1. The selection of these Lattice SBRT planning technique dimensions was based on previously published approaches, which used boost target vertices of 1-2 cm in diameter spaced 2-3 cm apart throughout the GTV ^11,12^. These vertices are defined within a physician-contoured GTV (GTV_2000), which included all visually identifiable gross disease. The GTV_2000 was expanded by 0.5–1.0 cm to create the PTV_2000, which was to receive 2000 cGy in 5 fractions. To generate the geometric lattice, the axial plane with the largest cross dimension within the GTV_2000 was first selected. Then, a 3 cm x 3 cm x 3 cm grid guide was overlaid on the GTV_2000, and the high-dose target vertices were placed at the grid intersections using a 1.5 cm diameter 3D brush to create the PTV_6670.

**Figure 1:**
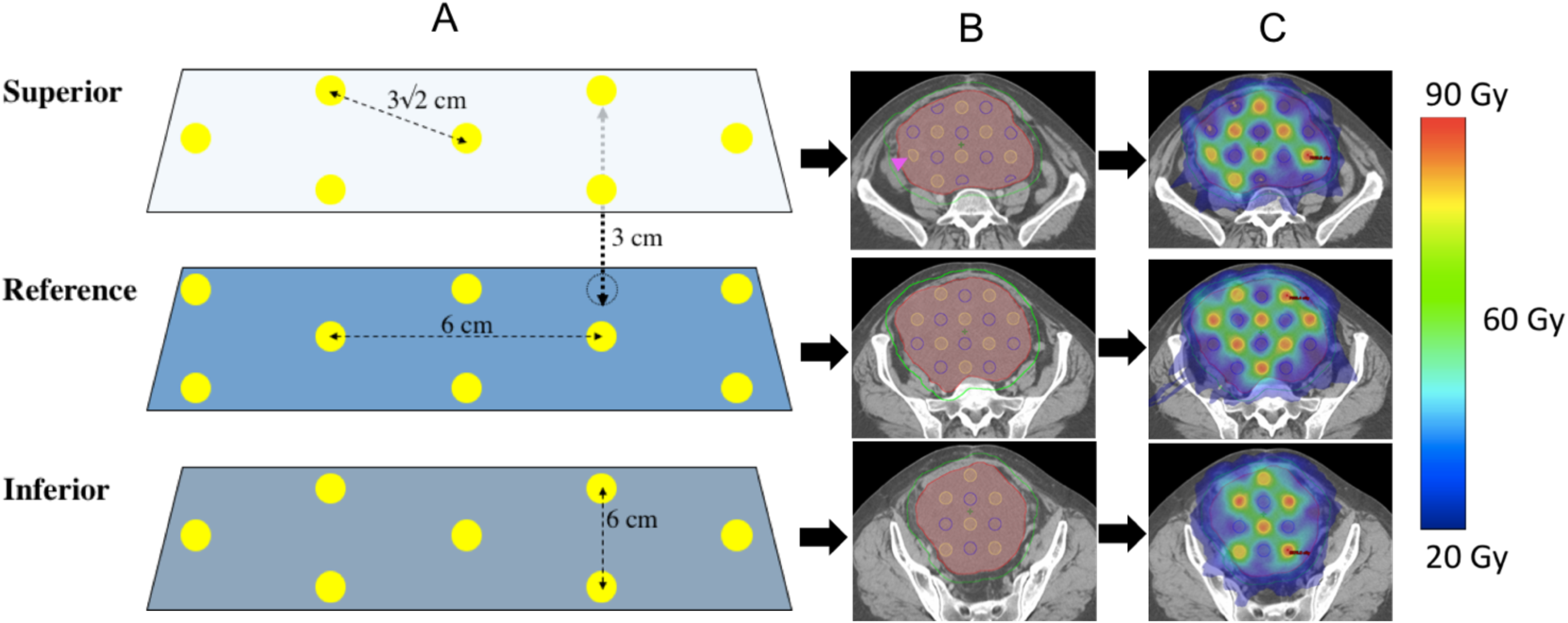
A) Geometric representation of sphere placement. Yellow dots represent the 1.5 cm diameter PTV_6670 target vertices, and dotted line vertices represent the transposed target vertices from adjacent planes. Axial planes where vertices are placed are separated by 3 cm in-plane. Within a plan, vertices are separated by 6 cm center to center (4.5 cm edge to edge) in orthogonal axes, and 3√2 cm along the diagonal. B) Axial CT slices of a target with the yellow outlined target vertices (PTV_6670) in each plane, red GTV_2000, and green PTV_2000. Magenta arrows denote cropped vertices in PTV_6670 that extend outside of the GTV_2000. C) Dose distribution after VMAT planning for the target with blue representing 20 Gy and red representing 66.7 Gy.

The PTV_6670 high-dose target vertices were alternated with 1.5 cm diameter avoidance vertices (i.e. PTV_Avoid) such that the center of a PTV_6670 vertex was 3 cm apart from a PTV_Avoid vertex. This process was repeated every 3 cm in the superior-inferior direction with vertices offset by 3.0 cm with respect to axial slices above and below. All PTV_6670 vertices extending outside of a 5 mm contraction of the GTV_2000 were completely removed to minimize spill of the 30 Gy isodose volume outside the PTV_2000. Additionally, any PTV_6670 vertices located within 1.5 cm of an OAR were completely removed to limit dose to normal tissue given any uncertainties at time of treatment. Finally, a PTV_Control structure was made by expanding PTV_6670 by 8 mm and subtracting this from PTV_2000, and was used to assess dose fall-off between adjacent PTV_6670 vertices.

The Lattice plan generation attempted to achieve a goal of ≥ 95% prescription dose coverage to at least 95% of both the PTV_2000 and PTV_6670. To achieve the high-dose gradients, each of the interspaced PTV_Avoid vertices required a minimum dose of between 19–20 Gy. High max point doses were allowed within the PTV_6670 vertices, with a maximum dose limited to 80 Gy. OAR constraints consistent with 5-fraction SBRT published in AAPM TG-101 were used in planning directives ^13(p101)^.

VMAT was used to achieve the internal high dose gradients, target coverages, and OAR objectives. VMAT plans are hypothesized to offer superior target coverage, reduce high-dose spill, and better spare OARs as compared with 3D conformal radiotherapy (3DCRT) for Lattice radiotherapy ^14,15^. After an initial attempt to generate both a 3DCRT and VMAT comparison plans for Lattice SBRT, we did not pursue 3DCRT plans further as it became clear that, regardless of the number of beam angles, we would not achieve OAR dose constraints while attaining our desired dose gradient. All plans were delivered on a Varian Truebeam using standard Millennium 120 MLC (5 mm/10 mm MLC widths). Once the Lattice SBRT contouring and treatment planning process was finalized, planning was completed for all 12 tumors utilizing Varian Eclipse treatment planning system v15.6, (Varian Medical Systems, Palo Alto, CA) using patient CT simulation scans.

After treatment planning, physician and physicist review, and plan approval, plan integrity and deliverability was evaluated following the standard clinical SBRT QA protocol. This included 2D EPID portal dosimetry to measure fluence throughout each angle of the beam’s arc, 1D IC absolute dose measurements within the PTV–6670 and PTV_AVOID (low-dose) vertices, and machine parameter delivery verification utilizing an in-house log-file program called Dyna QA ^16^. For EPID portal dosimetry, 2-dimensional fluence maps were measured for each arc and evaluated against the calculated fluence using a 3%/3mm criteria (95% pass rate) and 2%/2mm criteria (90% pass rate) across all pixels. The 2D planar measurements provided evidence of the accuracy of the high dose gradients within the GTV_2000. The 1D ion chamber measurements were completed using an small field Exradin A16 Ion Chamber (Standard Imaging, Middleton, WI) placed in an in-house designed solid water phantom at locations corresponding to the PTV_6670 and PTV_Avoid vertices, with one measurement in each structure captured per patient. A Dyna QA report was generated from the treatment machine log files and analyzed in-house software (MLC movement, gantry rotation, MUs delivered, etc.) ^16^.

## Results

The 11 patients had tumors of various histologies that were located in a range of anatomical locations (Table 1). Tumors had a median volume of 687.5 cc (range: 350-4440) and a median greatest axial dimension of 12.75 cm (range: 10-18.5). VMAT plans were created using flattening filter-free beams, multiple full or partial non-coplanar arcs with couch kicks up to 15° based on available clearance of the treatment couch. Plans used 6MV or 10MV energies with 10MV plans overall resulting in less monitor units (MU). A collimator rotation of 15°-90° and jaw tracking were used for all plans. Treatment delivery times were acquired during the IC QA process and ranged from 9–16 minutes (mean = 12.3 minutes), inclusive of couch kicks but exclusive of patient setup, imaging, or alignment.

**Table 1:** patient tumor characteristics Patient tumor characteristics for Lattice SBRT dosimetric and QA analysis ranked from smallest to largest volume.

| Target | Histology | Site | GTV (cc) | Largest axial dimension (cm) |
| --- | --- | --- | --- | --- |
| 1 | Cervical Squamous Cell Carcinoma | Central Pelvis | 350 | 10 |
| 2 | Leiomyosarcoma | Right Abdomen | 430 | 10.5 |
| 3 | Cholangiocarcinoma | Right Abdomen | 450 | 10 |
| 4 | Melanoma | Mediastinum | 490 | 13 |
| 5* | Undifferentiated sarcoma | Left Lung | 450 | 12.5 |
| 6* | Undifferentiated sarcoma | Right Neck | 615 | 12 |
| 7 | Leiomyosarcoma | Left Lung | 745 | 12.5 |
| 8 | Liposarcoma | Central Pelvis | 1805 | 13 |
| 9 | Sarcomatoid carcinoma | Right Abdomen | 940 | 13.5 |
| 10 | Liposarcoma | Right Pelvis | 1035 | 14.5 |
| 11 | Melanoma | Left Abdomen/Pelvis | 1600 | 15.5 |
| 12 | Malignant Peripheral Nerve Sheath Tumor | Left Abdomen/Pelvis | 4475 | 18.5 |

All plans, except one, met dose constraints for OARs (Supplementary Table 1). The skin dose constraint could not be achieved in this patient who had a very large sarcoma metastasis to the neck where the tumor extended close to the skin. All targets, achieved > 95% coverage for the PTV_2000 and PTV_6670 (Table 2). Target 12, the largest target, was able to achieve adequate dose coverage and OAR sparing, however, the DMean of the PTV_Avoid was the highest of this series, indicating that it may be difficult to achieve desired dose fall off in exceptionally large targets. An additional metric, termed the *Lattice composite*, was defined as PTV_6670 divided by GTV_2000 to represent the volume of tumor target filled by high-dose vertices. All patients had a Lattice composite of approximately 2-4%.

**Table 2:** target dose coverage, Lattice Composite 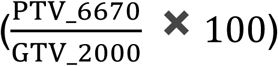, number of non-coplanar arcs, MU per Gy for each treatment plan, Selectivity 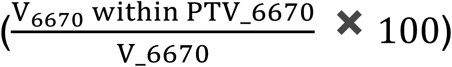 and Gradient Index of the PTV_6670 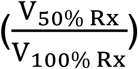

| Target | Site | GTV<br>(cc) | V95%Rx |  | PTV_Avoid<br>DMean (Gy) | Lattice<br>Composite | #<br>Arcs | MU per<br>cGy | Selectivity | Gradient<br>Index |
| --- | --- | --- | --- | --- | --- | --- | --- | --- | --- | --- |
|  |  |  | PTV_6670 | PTV_2000 |  |  |  |  |  |  |
| 1 | Central Pelvis | 350 | 99.9 | 100 | 21.8 | 3.1% | 4 | 2.18 | 84.3 | 16.7 |
| 2 | Right Abdomen | 430 | 99.8 | 100 | 26.7 | 2.3% | 4 | 3.63 | 67.7 | 14.5 |
| 3 | Right Abdomen | 450 | 100 | 100 | 21.0 | 2.6% | 4 | 3.03 | 60.7 | 22.2 |
| 4 | Mediastinum | 490 | 100 | 100 | 21.3 | 1.9% | 5 | 3.29 | 59.2 | 19.1 |
| 5 | Left Lung | 450 | 99.5 | 99.7 | 20.8 | 4.3% | 4 | 4.7 | 65.7 | 14.5 |
| 6 | Right Neck | 615 | 89.6 | 96.5 | 22.2 | 3.1% | 3 | 3.31 | 60.0 | 14.4 |
| 7 | Left Lung | 745 | 95.3 | 100 | 21.6 | 2.2% | 4 | 2.88 | 74.1 | 23.9 |
| 8 | Central Pelvis | 1805 | 99.1 | 99.9 | 25.8 | 2.2% | 4 | 4.87 | 86.2 | 24.4 |
| 9 | Right Abdomen | 940 | 99.8 | 100 | 22.4 | 2.3% | 4 | 5.48 | 67.0 | 11.2 |
| 10 | Right Pelvis | 1035 | 97.2 | 99.4 | 22.2 | 2.3% | 4 | 3.76 | 79.0 | 26.2 |

All 2D EPID, IC, and Dyna QA results achieved the thresholds specified, as specified in Supplementary Table 2. The IC measurements, taken within the PTV_6670 structures, were within 3% of the expected dose predicted by the TPS. A larger deviation of agreement was observed for the low-dose IC measurements taken in the PTV_Avoid structure, with no measurement exceeding 5% deviation.

## Discussion

This study demonstrates that the proposed Lattice SBRT technique adheres to established SBRT safety guidelines, and can be planned and delivered on standard commercially available equipment. Lattice SBRT plans were successfully created for tumors ranging from 350-4440 cc, and the plans met tumor coverage objectives and OAR dose constraints for 5-fraction SBRT. The planning procedure was critical to consistently achieve the high dose gradients characteristic of Lattice planning. The 1.5 cm diameter of PTV_6670 and PTV_Avoid vertices was selected for three reasons. First, it represented a volume that approached the smallest limit that could be accurately, consistently, and safely delivered across multiple off-axis locations within the larger GTV_2000 tumor using a standard MLC-based delivery. Second, it provided ample margin to minimize the impact of dose smearing caused by daily setup variations, thus maintaining the desired 100% - 30% dose fall-off. Third, the 1.5 cm spacing between the PTV_6670 and PTV_Avoid vertices represented the minimum achievable distance needed to create the desired dose fall-off (Supplemental Figure S1). Furthermore, the overall Lattice structure was critical in defining desired fluence paths to reach internal PTV_6670 vertices without increasing dose to the PTV_Avoid vertices (Supplemental Figure S2). While the described planning method isn’t the exclusive means to create Lattice SBRT plans, it is a consistent procedure that can be applied successfully across a broad range of tumor volumes, sites, and clinics.

All plans passed QA testing following a SBRT QA protocol with EPID portal dosimetry at 3%/3mm and 2%/2mm, IC absolute dose measurements at high- and low-dose points, and Dyna QA to confirm mechanical deliverability of plans. The increased variability in the low-dose IC measurements is attributed to the higher inaccuracy of the inter- and intra-MLC modeling and the associated length of time that low-dose regions are covered under the MLCs during the treatment process. It should be noted that the reported values are relative to the expected local dose produced in the TPS, resulting in a deviation of ∼5% being equivalent to 1 Gy in the low-dose measurements. This ∼5% local deviation in the low dose region corresponds to ∼1% - 2% of the global prescription and is within the 2% tolerance for IC measurements as specified in AAPM Task Group 218 ^17^. Both the IC and EPID-based measurements followed PSQA recommendations provided in AAPM Task Group 218. As the EPID-based measurements are inherently perpendicular to the delivery direction, the full arc-by-arc fluence distribution is evaluated against the corresponding distribution produced in the planning system.

There were several limitations inherent to the retrospective nature of the study. First, all patients were clinical treated following a palliative regime. As such, the optimal patient positioning and immobilization for SBRT treatments wasn’t always present. For example, in two lung cases, the patient’s arms were positioned down and were directly in the beam path for all arcs. To maintain the standard lung SBRT positioning, the arms were subsequently overridden to air. Additionally, many of the patients didn’t have immobilization common for SBRT treatments, including abdominal compression and body length immobilization bags. However, these limitations would impact the delivery of the Lattice plans and not the planning procedure described in this study. Standard SBRT positioning, immobilization, and on treatment imaging practices should be followed for any patients treated using the lattice procedure described in this study.

We have recently completed evaluation of the safety of this regimen in a phase I clinical trial for patients with large, unresectable tumors (NCTXXXX) and are now evaluating its efficacy for patients with specific histologies (NCTXXXX). Future work will evaluate delivering Lattice SBRT using intensity modulated proton therapy and magnetic resonance guided adaptive radiotherapy platforms.

## Conclusion

Lattice SBRT is an approach to deliver dose-escalated radiation to large tumors in a way that may surmount the limitations of conventional fractionation. Our approach utilizes VMAT to deliver high-dose “islands” within a “sea” of lower dose. Our approach is clinically and technically feasible, and is being prospectively evaluated in two ongoing trials for patients with large, unresectable tumors.

## Data Availability

The data referred to in this manuscript is not available.

## Contouring, Treatment Planning, and Evaluation Procedure for Lattice SBRT

### Contouring

1. The physician contours the GTV_2000, which represents the gross tumor volume. The GTV_2000 is expanded 0.5-1.0 cm to form the PTV_2000.
2. Contour all normal structures in their entirety. High density and machine couch contours should be included where appropriate.
3. Create the PTV_6670 and PTV_Avoid structures.
4. Turn on the grid in the contouring workspace and set to user-defined 3.0 cm.
5. Locate the center of PTV_2000 and set the viewing planes to match the grid intersection in all 3 planes.
6. Select PTV_6670 and select a 1.5 cm 3D brush.
7. Working in the axial plane, draw PTV_6670 spheres at every other grid intersection point, then repeat the process with PTV_Avoid spheres. The spheres should encompass and surround the PTV in this plane. PTV_6670 and PTV_Avoid spheres can be deleted and/or rearranged later if needed.
8. Move one plane superior to the next grid intersection point and place PTV_6670 and PTV_Avoid spheres so that they alternate in the superior-inferior direction.
9. Continue this alternating pattern until the superior most region of PTV_2000 is covered
10. Repeat the process in the inferior direction so that the entire PTV_2000 is covered.
11. Finalize the spheres by translating the structures and/or switch the PTV_6670 and PTV_Avoid to optimize sphere placement.
12. Create a PTV_Control structure by cropping PTV_6670 + 8 mm from PTV_2000.
13. Crop PTV_6670 so that they do not go outside the GTV_2000 – 5 mm volume.
14. Crop PTV_Avoid so that they do not go outsdie of the PTV_2000.
15. Ensure PTV_6670 spheres are not overlapping any critical OAR plus some volume.

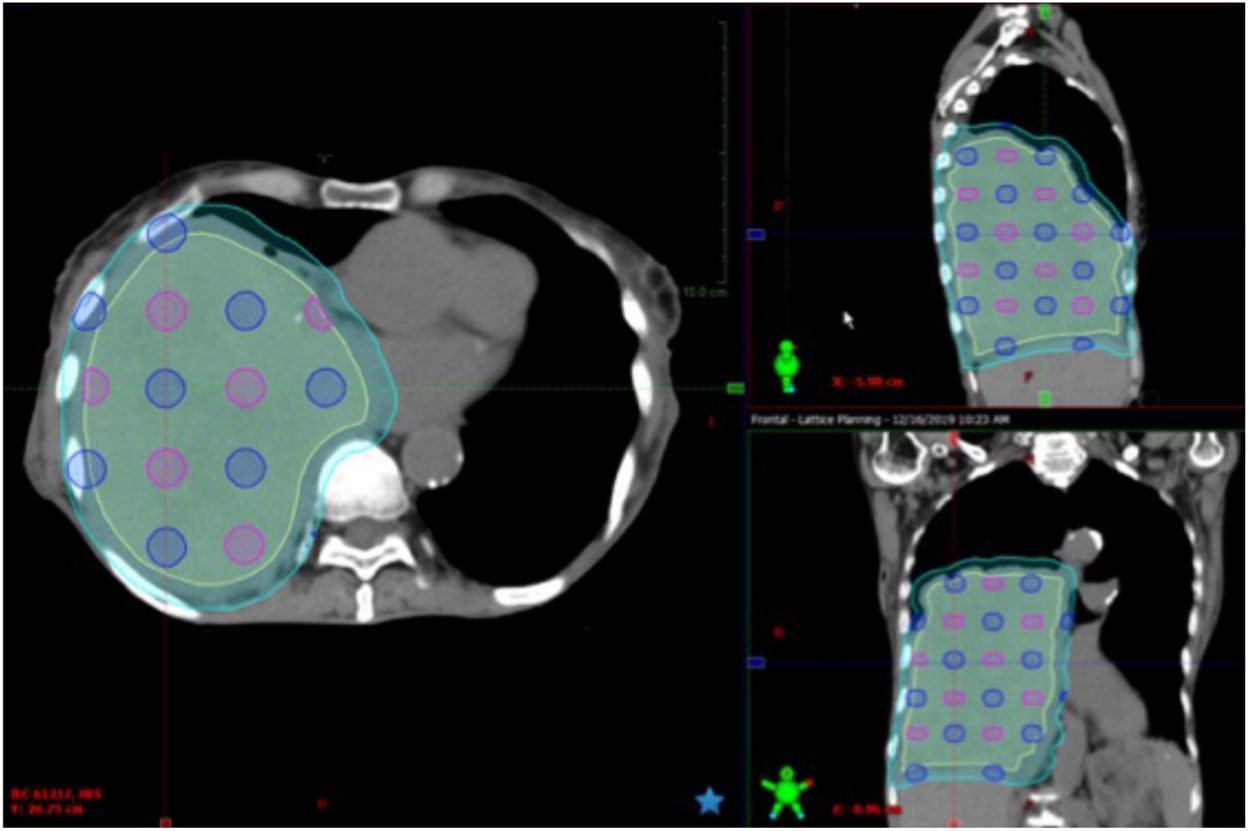

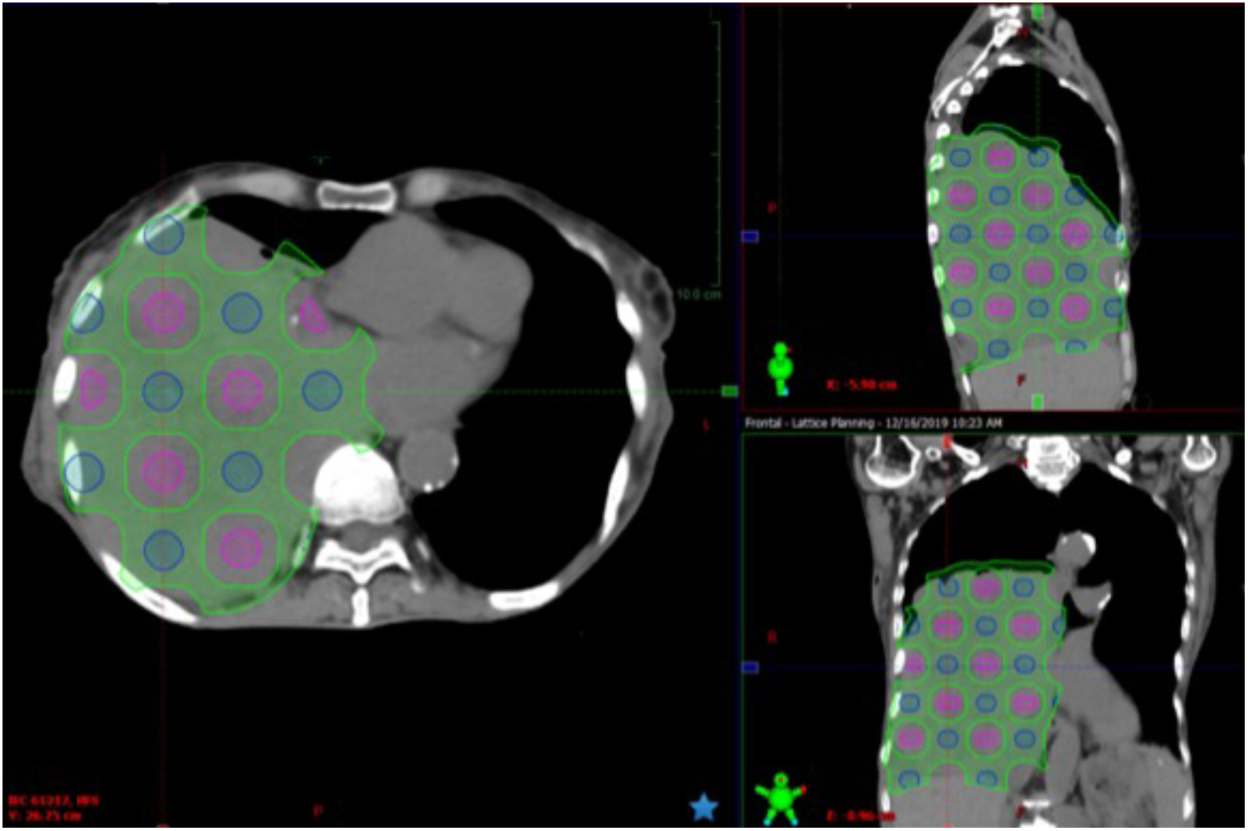

### Planning and Optimization

*Plans are created for the Varian Truebeam or Edge using an SRS ARC technique at our institution. The following is a suggested planning process to cover the target volumes designed above. Suggested optimization parameters are included below*.

1. Insert a new course and plan with a prescription of 6670 cGy in 5 fractions. PTV_6670 is the planning target and reference point^1^. Ensure the isocenter is in the center of PTV_2000.
2. Select 6MV or 10MV depending on the treatment site (10MV may result in lower overall monitor units).
3. Utilize full or partial arcs to plan the treatment. Most volumes are sufficiently covered with 4 arcs. Couch kicks up to 10° may be used.
4. Jaw tracking with initial jaws set to PTV_2000 and a collimator rotation of 15°-90° should be used.
5. Typical MU objectives and baseline normal tissue objectives are shown. Normal tissue objectives are set for rapid dose fall off.
6. OAR objectives should be given a high optimization priority.
7. Pause at each MR level accordingly to ensure optimal plan solution if found.

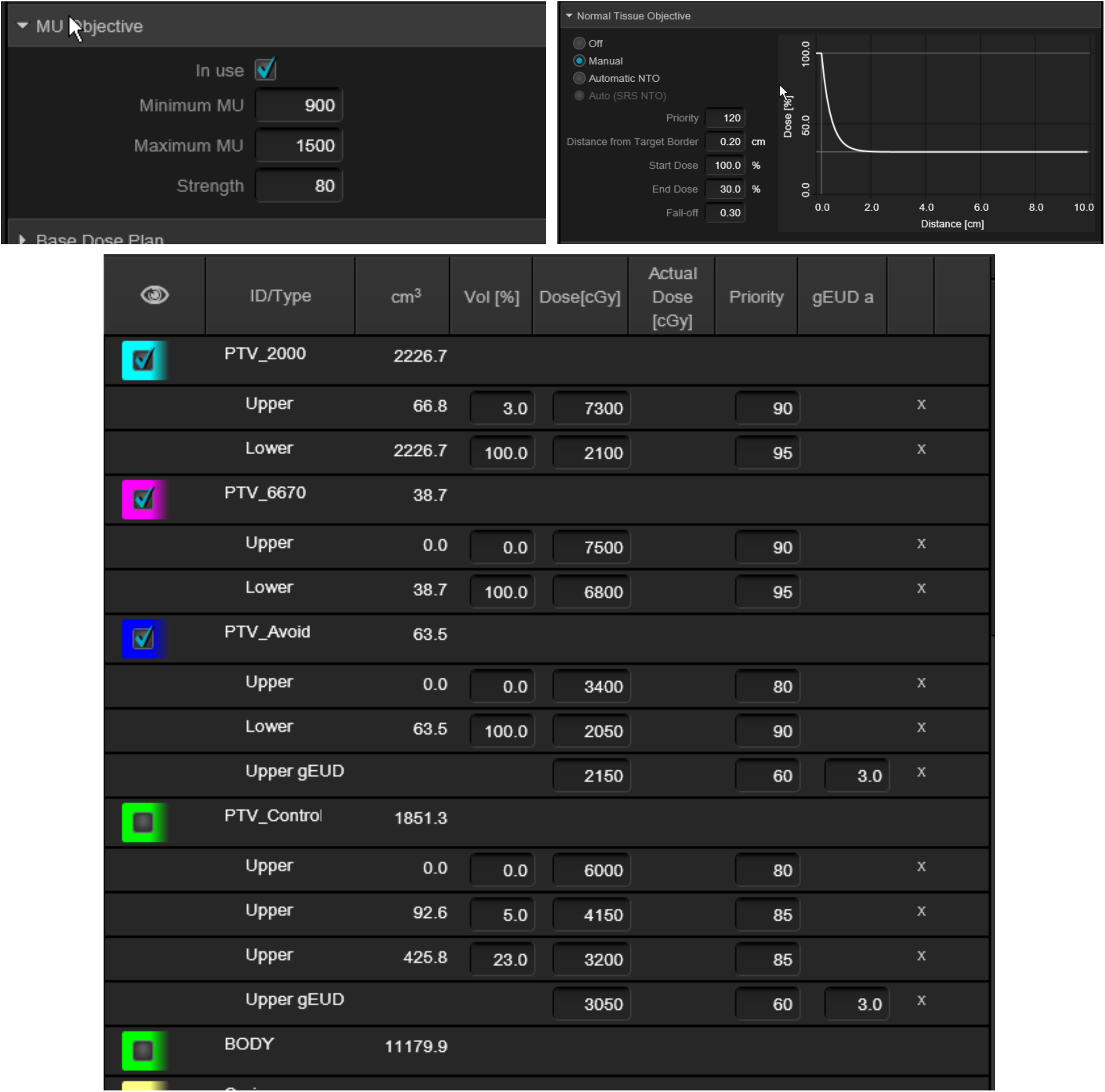

### Evaluation

1. Clinical OAR goals are the top priority. They cannot be exceeded in our current clinical trials. PTV_6670 coverage is a secondary goal for evaluation. If a PTV_6670 sphere proximal to an OAR is causing the plan to exceed the OAR goal, that sphere can be retracted or deleted.
2. Once OAR goals are met, a subjective analysis should be performed to evaluate the following features:
  a. Overall lattice distribution with isodose holes at 2400 cGy and below present between PTV_6670 spheres.
  b. Global hot spot approximately 115%.
  c. PTV_Avoid structure should have a mean dose between 2000-2400 cGy.
  d. PTV_2000 and PTV_6670 should have V95% Rx > 95%. However, under coverage of targets is allowed on current clinical trials in order to meet dose constraints.
  e. MU ratio between 2.0 and 3.0.
3. The beam parameters for an example plan are shown on the next page.

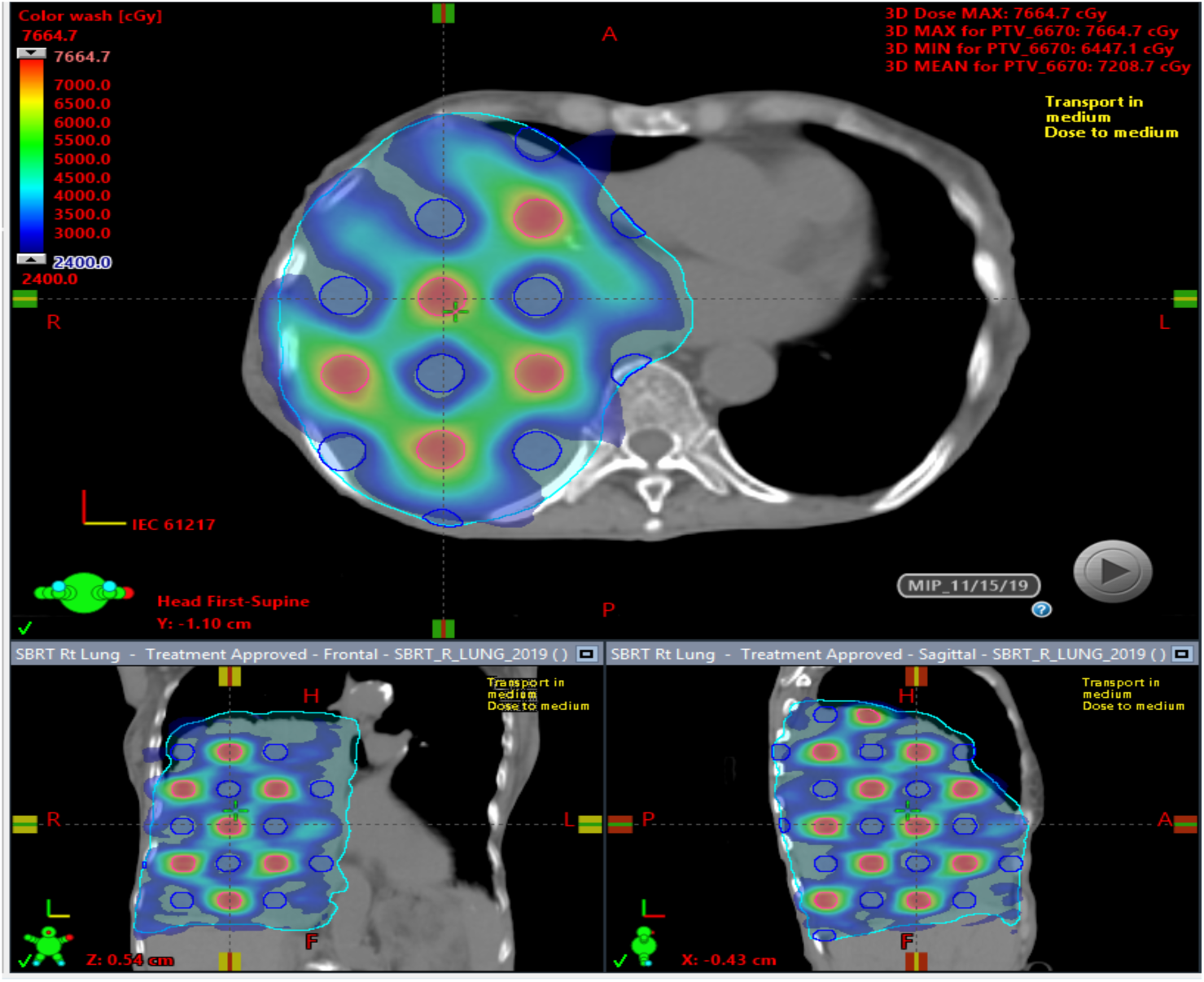

### Troubleshooting and Tips

1. Evaluate the PTV_6670 and PTV_Avoid spheres and beam arrangement with a physicist colleague before optimization. Beam selection will be disease site specific.
2. Don’t directly optimize on the mean dose to PTV_Avoid, as this seems to lead to poor PTV_2000 target coverage.
3. Be sure to check for beam clearance with a physicist or therapist.
4. Ensure the correct table is inserted if required.
5. Have fun!

## Beam Parameters

| # | 1 | 2 | 3 | 4 | 5 | 6 | 7 | 8 | 9 |
| --- | --- | --- | --- | --- | --- | --- | --- | --- | --- |
| Type | Rapid Arc I<br>MRT | Rapid Arc I<br>MRT | Rapid Arc I<br>MRT | Rapid Arc I<br>MRT | Setup Bea<br>m | Setup Bea<br>m | Setup Bea<br>m | Setup Bea<br>m | Setup Bea<br>m |
| Field ID | *1A | *1B | *1C | *1D | CBCT | APSU | RTSU | PASU | LTSU |
| Field Name | 181 CW 45 | 45 CCW 1<br>81 | 181 CW 45 | 45 CCW 1<br>81 | N/A | N/A | N/A | N/A | N/A |
| Is Setup Beam | No | No | No | No | Yes | Yes | Yes | Yes | Yes |
| Machine | BJH_TR7 |  |  |  |  |  |  |  |  |
| Scale | IEC61217 |  |  |  |  |  |  |  |  |
| Modality | Photon |  |  |  |  |  |  |  |  |
| Energy | 10X | 10X | 10X | 10X | 6X | 6X | 6X | 6X | 6X |
| Gantry Angle | 181.0 CW<br>45.0 | 45.0 CCW<br>181.0 | 181.0 CW<br>45.0 | 45.0 CCW<br>181.0 | 0.0 | 0.0 | 270.0 | 180.0 | 90.0 |
| Couch Angle | 0.0 | 0.0 | 10.0 | 350.0 | 0.0 | 0.0 | 0.0 | 0.0 | 0.0 |
| Isocenter | (0.00, 0.00, 0.00) |  |  |  |  |  |  |  |  |
| Collimator Angle | 15.0 | 345.0 | 45.0 | 315.0 | 0.0 | 0.0 | 0.0 | 0.0 | 0.0 |
| X1 [cm] | -8.00 | -7.00 | -8.00 | -7.00 | -10.00 | -10.00 | -10.00 | -10.00 | -10.00 |
| X2 [cm] | 7.00 | 7.76 | 6.93 | 8.00 | 10.00 | 10.00 | 10.00 | 10.00 | 10.00 |
| Field X [cm] | 15.00 | 14.76 | 14.93 | 15.00 | 20.00 | 20.00 | 20.00 | 20.00 | 20.00 |
| Y1 [cm] | -12.00 | -12.00 | -12.00 | -13.00 | -10.00 | -10.00 | -10.00 | -10.00 | -10.00 |
| Y2 [cm] | 12.00 | 11.00 | 12.00 | 12.00 | 10.00 | 10.00 | 10.00 | 10.00 | 10.00 |
| Field Y [cm] | 24.00 | 23.00 | 24.00 | 25.00 | 20.00 | 20.00 | 20.00 | 20.00 | 20.00 |
| Dose Rate [MU/min] | 600 | 600 | 600 | 600 | 600 | 600 | 600 | 600 | 600 |
| SSD [cm] | 91.10 | 87.56 | 91.09 | 87.76 | 91.16 | 91.16 | 93.66 | 91.08 | 82.11 |
| MU | 819.73 | 848.51 | 901.43 | 738.04 | 0.00 | 0.00 | 0.00 | 0.00 | 0.00 |
| Total MU | 3307.70 |  |  |  |  |  |  |  |  |
| MU Ratio | 2.48 |  |  |  |  |  |  |  |  |
| Complexity | 0.272 | 0.269 | 0.208 | 0.209 | N/A | N/A | N/A | N/A | N/A |
| Leaf Motion<br>Calculator | N/A | N/A | N/A | N/A | N/A | N/A | N/A | N/A | N/A |
| Number of Control<br>Points | 114 | 114 | 114 | 114 | 2 | 2 | 2 | 2 | 2 |
| Total Control Points | 466 |  |  |  |  |  |  |  |  |
| Applicator | No | No | No | No | No | No | No | No | No |
| Bolus | No | No | No | No | No | No | No | No | No |
| Block | No | No | No | No | No | No | No | No | No |
| Wedge | No | No | No | No | No | No | No | No | No |
| Compensator | No | No | No | No | No | No | No | No | No |
| Use MLC | Yes | Yes | Yes | Yes | No | No | No | No | No |
| Tray | No | No | No | No | No | No | No | No | No |
| 1A+ |  |  |  |  |  |  |  |  |  |

**Supplementary Table 1:**
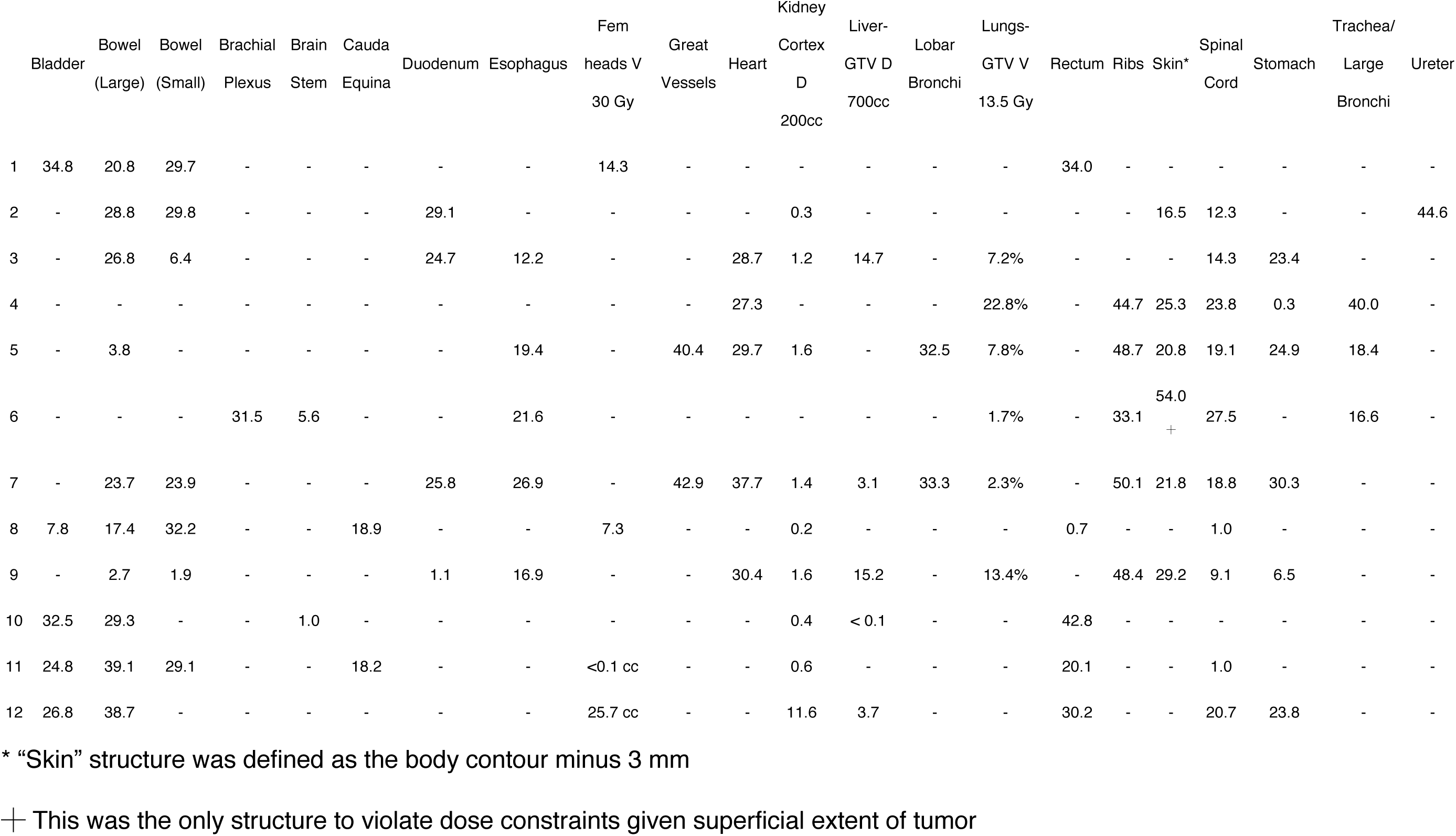
Dose to OARs for each tumor treatment plan.

|  | Bladder | Bowel<br>(Large) | Bowel<br>(Small) | Brachial<br>Plexus | Brain<br>Stem | Cauda<br>Equina | Duodenum | Esophagus | Fem<br>heads V<br>30 Gy | Great<br>Vessels | Heart | Kidney<br>Cortex<br>D<br>200cc | Liver-GTV D<br>700cc | Lobar<br>Bronchi | Lungs-GTV V<br>13.5 Gy | Rectum | Ribs | Skin* | Spinal<br>Cord | Stomach | Trachea/<br>Large<br>Bronchi | Ureter |
| --- | --- | --- | --- | --- | --- | --- | --- | --- | --- | --- | --- | --- | --- | --- | --- | --- | --- | --- | --- | --- | --- | --- |
| 1 | 34.8 | 20.8 | 29.7 | - | - | - | - | - | 14.3 | - | - | - | - | - | - | 34.0 | - | - | - | - | - | - |
| 2 | - | 28.8 | 29.8 | - | - | - | 29.1 | - | - | - | - | 0.3 | - | - | - | - | - | 16.5 | 12.3 | - | - | 44.6 |
| 3 | - | 26.8 | 6.4 | - | - | - | 24.7 | 12.2 | - | - | 28.7 | 1.2 | 14.7 | - | 7.2% | - | - | - | 14.3 | 23.4 | - | - |
| 4 | - | - | - | - | - | - | - | - | - | - | 27.3 | - | - | - | 22.8% | - | 44.7 | 25.3 | 23.8 | 0.3 | 40.0 | - |
| 5 | - | 3.8 | - | - | - | - | - | 19.4 | - | 40.4 | 29.7 | 1.6 | - | 32.5 | 7.8% | - | 48.7 | 20.8 | 19.1 | 24.9 | 18.4 | - |
| 6 | - | - | - | 31.5 | 5.6 | - | - | 21.6 | - | - | - | - | - | - | 1.7% | - | 33.1 | 54.0<br>+ | 27.5 | - | 16.6 | - |
| 7 | - | 23.7 | 23.9 | - | - | - | 25.8 | 26.9 | - | 42.9 | 37.7 | 1.4 | 3.1 | 33.3 | 2.3% | - | 50.1 | 21.8 | 18.8 | 30.3 | - | - |
| 8 | 7.8 | 17.4 | 32.2 | - | - | 18.9 | - | - | 7.3 | - | - | 0.2 | - | - | - | 0.7 | - | - | 1.0 | - | - | - |
| 9 | - | 2.7 | 1.9 | - | - | - | 1.1 | 16.9 | - | - | 30.4 | 1.6 | 15.2 | - | 13.4% | - | 48.4 | 29.2 | 9.1 | 6.5 | - | - |
| 10 | 32.5 | 29.3 | - | - | 1.0 | - | - | - | - | - | - | 0.4 | < 0.1 | - | - | 42.8 | - | - | - | - | - | - |
| 11 | 24.8 | 39.1 | 29.1 | - | - | 18.2 | - | - | <0.1 cc | - | - | 0.6 | - | - | - | 20.1 | - | - | 1.0 | - | - | - |
| 12 | 26.8 | 38.7 | - | - | - | - | - | - | 25.7 cc | - | - | 11.6 | 3.7 | - | - | 30.2 | - | - | 20.7 | 23.8 | - | - |

\* "Skin" structure was defined as the body contour minus 3 mm
+ This was the only structure to violate dose constraints given superficial extent of tumor

**Supplementary Table 2:** QA results for each patient.

| Target | Site | GTV (cc) | Portal Dosimetry (%) |  | Ion Chamber* (%) |  | Dyna QA |  |
| --- | --- | --- | --- | --- | --- | --- | --- | --- |
|  |  |  | 2mm/2% | 3mm/3% | PTV_6670 | PTV_Avoid | 2mm/2% | 3mm/3% |
| 1 | Central Pelvis | 350 | 99.9 | 1000 | 99.3 | 102.1 | 100 | 100 |
| 2 | Right Abdomen | 430 | 99.7 | 99.8 | 100.1 | 102.1 | 100 | 100 |
| 3 | Right Abdomen | 450 | 99.9 | 100 | 99.1 | 101.3 | 100 | 100 |
| 4 | Mediastinum | 490 | 95.3 | 98.9 | 98.7 | 100.0 | 100 | 100 |
| 5* | Left Lung | 450 | 99.7 | 100 | 101.1 | 105.0 | 100 | 100 |
| 6* | Right Neck | 615 | 82.3 | 96.9 | 100.0 | 103.8 | 100 | 100 |
| 7 | Left Lung | 745 | 99 | 99.9 | 99.9 | 101.5 | 100 | 100 |
| 8 | Central Pelvis | 1805 | 99.2 | 99.9 | 98.9 | 100.2 | 100 | 100 |
| 9 | Right Abdomen | 940 | 97.2 | 99.7 | 99.3 | 103.0 | 99.7 | 100 |
| 10 | Right Pelvis | 1035 | 99.9 | 100 | 99.9 | 101.9 | 100 | 100 |
| 11 | Left Abdomen/Pelvis | 1600 | 98.8 | 99.9 | 98.8 | 104.5 | 100 | 100 |
| 12 | Left Abdomen/Pelvis | 4475 | 97.0 | 99.6 | 99.4 | 101.2 | 100 | 100 |

\* Calculated as dose measurement during QA divided by goal dose

**Figure S1:**
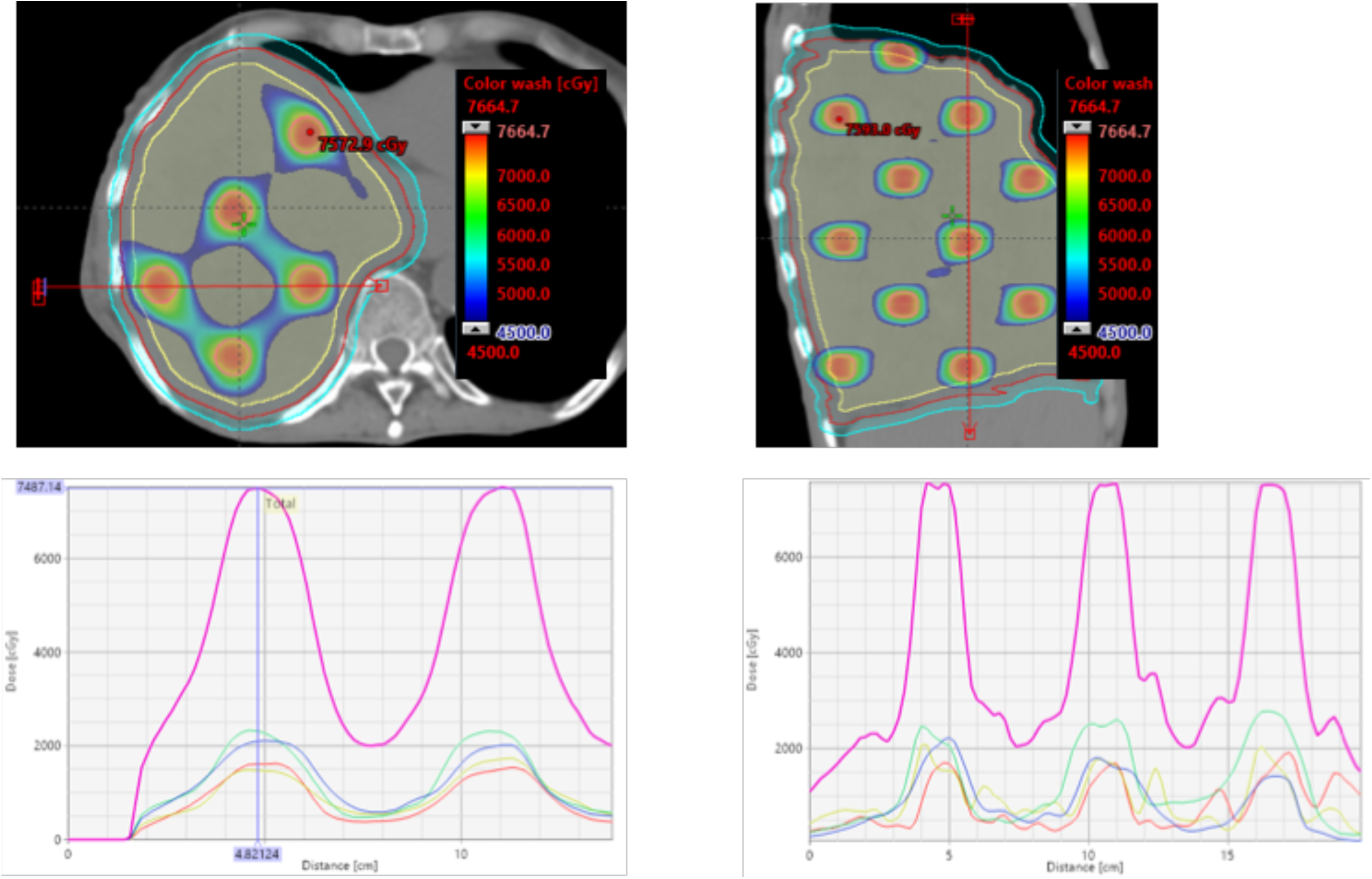
Dose profiles illustrating the high dose gradients in the A) axial and B) cranial-caudal directions. Note the dose gradient in the axial plane produces a dose gradient ranging from 66.7Gy to 25Gy over 1.5cm. This dose gradient is steeper in the cranial-caudal direction.

**Figure S2:**
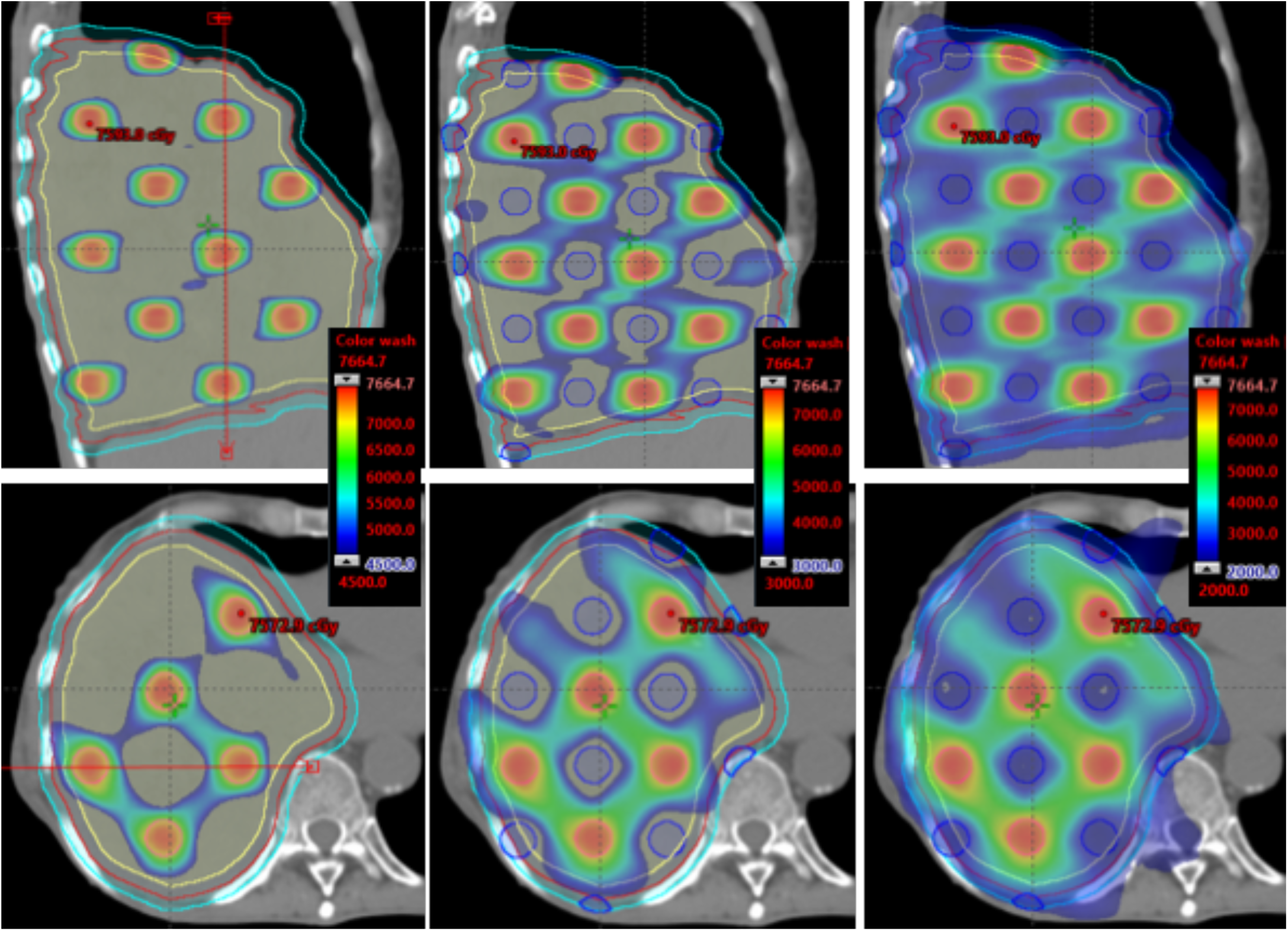
Dosimetric structure produce following the Lattice SBRT method, illustrated in the cranial-caudal (upper) and axial (lower) planes. The specific structure creates preferred fluence paths, visible in the diagonal directions, allowing the creation of the high dose gradients in the interior of the large tumors treated with this technique.

## Footnotes

1 Plans were created for Varian Truebeam or Edge in this study.

